# Pregnancy intentions in the Netherlands: An evaluation of a multidimensional and continuous construct

**DOI:** 10.1101/2023.11.13.23298453

**Authors:** Merel Sprenger, Wieke Y. Beumer, Jenneke van Ditzhuijzen, Jessica C. Kiefte-de Jong

## Abstract

**Background:** Pregnancy intention is a multidimensional, dynamic as well as context-specific construct. The primary aim of this study is to establish a reliable survey on pregnancy intention in the Netherlands based on the London Measure of Unplanned Pregnancy (LMUP).

**Methodology:** The development of the Dutch Adaptation of the London Measure of Unplanned Pregnancy (DA-LMUP) was based on the LMUP, but substantial differences were made, resulting in an adapted version both for people who were pregnant and one for partners. We conducted a psychometric evaluation of the DA-LMUP using observational data from two studies in the Netherlands, with 1172 participants (both pregnant people and their partners), aged between 16-55 years. Next, acceptability and reliability, including test-retest stability were assessed. Further, construct and convergent validity were analysed – including hypothesis testing – and Mokken scaling was applied.

**Results:** The DA-LMUP demonstrated to be acceptable and reliable (Cronbach’s alpha >0.80 for both versions: pregnant people and partners). Construct validity was acceptable and Mokken scale analyses indicated a strong scale (H-coefficient: 0.68). All hypothesis tests were confirmed: people who were younger, single, and nulliparous reported lower pregnancy intent. People who had an abortion reported lower pregnancy intent compared to people continuing their pregnancy to term. Lastly, results showed a high correlation of pregnancy intent in couples (n = 257 couples).

**Conclusions:** The DA-LMUP is evaluated as reliable for the Dutch context. It offers researchers a suitable instrument to measure pregnancy intention in a multidimensional manner, constituting a closer reflection of people’s actual experiences.

## 1 Introduction

A positive pregnancy test elicits a wide variety of emotional responses. Although unintended pregnancy is a common phenomenon worldwide, it is often considered a public health issue (1). In most research and policy, a pregnancy is defined as either wanted, mistimed (unplanned, but wanted) or unwanted (1, 2). Defining pregnancy intentions in this categorical manner leads to oversimplification(2-5). People often experience ambivalent attitudes towards (the prospect of) pregnancy (6). Some people feel happy at the prospect of a pregnancy, while simultaneously wanting to prevent conception (7). Moreover, often people don’t have fully formed family building plans (yet) (8).

To address pregnancy intentions in a manner that is closer to people’s actual experiences in research, calls have been made for using a multidimensional measurement on a continuous scale (9). The London Measure of Unplanned Pregnancy (LMUP) – assessed as easy to understand, valid and reliable – is a retrospective instrument that can be used for this purpose (10). The LMUP was developed as a short, self-administered measure and has been translated and validated in several other languages across all continents (11). It considers three key dimensions: 1) context (timing and partner discussion), 2) stance (desire for pregnancy/parenthood and expressed intentions), 3) behaviour (contraceptive use and pre-conceptual preparations).

In the current study, we developed the Dutch Adaptation of the London Measure of Unplanned Pregnancy (DA-LMUP), based on the LMUP (10). We introduced substantial modifications to tailor the instrument to the Dutch context, making it unsuitable for direct comparison with the LMUP. Although a validated Flemish-Dutch version of the LMUP exists (12), Flemish and Dutch are distinct language varieties, making the Flemish-Dutch version insufficiently accessible for use in the Netherlands (13). Moreover, societal norms and perceptions surrounding unintended pregnancy can differ significantly across political, cultural, and healthcare systems, including systems in the Netherlands and Belgium (14, 15).

Furthermore, there is growing recognition of the importance of incorporating partners’ perspectives in research on reproductive health (16). Therefore, it is important to adapt and evaluate pregnancy intention measures tailored to partners. We define ‘partners’ as the people who are involved in the pregnancy either as a romantic partner (regardless of their gender and/or sex), or a non-romantic, but sexually or biologically involved partner. Up to now, the LMUP has been primarily tested in partners of people continuing their pregnancies to term, mostly with quite intended pregnancies (17, 18). Earlier research indicated that there is a high consistency of pregnancy intentions within couples, i.e., couples often report similar pregnancy intentions (19, 20), but recent evidence, and evidence in couples opting for abortion, is lacking.

This study aims to establish and evaluate two versions of DA-LMUP, applicable to pregnant people and their partners, for use in the Dutch context so that it might be used in Dutch monitor studies on sexual and reproductive health.

## 2 Methods

### 2.1 Developing the questionnaire

We based our first version of the DA-LMUP on the Dutch-Flemish LMUP and the UK 2020 update, incorporating linguistic changes from Flemish to Dutch (12, 21). Second, this version was evaluated in discussions with five other researchers, three professionals from the field (an abortion doctor, sexologist, and midwife), seven experience experts (of which five women) and with two linguists. After making adjustments based on their input, a blind back-translation into English was performed by two independent translators with English as their native language. This version was then compared to the original LMUP. No adjustments had to be made. Third, the surveys were evaluated in individual think-aloud interviews with five women and two partners who recently experienced an (unintended) pregnancy. Participants talked the interviewer through the cognitive process of completing the survey and expressed any thoughts or feelings that came up (22, 23). Based on these interviews, the questionnaire was slightly adjusted. Again, this version was discussed with the experts. Consensus was reached, resulting in the final version, which was reviewed in consultation with the developers of the LMUP, who provided guidance on next steps. They advised us to clearly document our adaptations, emphasising that the DA-LMUP, though based on the LMUP, has been substantially adapted to align with the Dutch context and should be regarded as a distinct instrument. All adjustments are presented in Table 1. The two DA-LMUP versions are available in the supplementary material.

**Table 1.** Adjustments made compared to the Flemish and UK 2020 LMUP versions.^a^.

| Question | Change | Reason |
| --- | --- | --- |
| 1 | Move of 'I/we were using' from beginning of answer option to end of question | To improve readability |
| 1 | Clarification of answer option on always using contraception, but knowing method had failed | To improve readability, based on think aloud interviews |
| 2 | Addition of 'parent' next to 'mother' | To improve acceptability |
| 2P | Use of 'father/parent' instead of 'mother/parent' | To improve acceptability |
| 3P | Use of 'having a child' instead of 'becoming pregnant' | To improve acceptability and to better suit Dutch language, based on think aloud interviews |
| 3P, 4P, 5P | Use of 'before the pregnancy' instead of 'before I got pregnant' | To improve acceptability and to better suit Dutch language, based on think aloud interviews |
| 4 | Use of word 'child' instead of 'baby' | To better suit Dutch language |
| 5 | Abbreviation of introductory text | To improve readability and acceptability |
| 5 | Addition of answer option in which participants agreed with their partner that they did not want any (more) children | To accommodate this group, based on think aloud interviews |
| 6 | Addition of answer option in which participants report exercising more | To prompt participants to think of this health behaviour |
| 6P | Remove option of folic acid | To improve acceptability |
| All | Addition of answer options 'I don't know' or 'I'd rather not say' | Necessary for our data collection as for all items, answers were required |

453 <sup>a</sup>Note: for question numbers followed by a 'P', changes were made in the version for partners only.

### 2.2 Data collection

#### 2.2.1 Design

We combined data from two Dutch prospective population-based cohort studies: the BluePrInt study and the RISE UP study. Both are part of a Dutch consortium around unintended pregnancy funded by ZonMw. Primary data collection took place from May 2022 to August 2023. Both studies were reviewed by the ethical committee and received a waiver from the Medical Research Involved Human subjects Act (WMO): the RISE UP study from the Medical Research Ethics Committee of Leiden Den Haag Delft under reference number N21.127, and the BluePrInt study from the Medical Research Ethics Committee of the Amsterdam Medical Center under reference number W21_407. All participants provided consent before participating.

#### 2.2.2 Participants

The BluePrInt study (n = 910) included people who carried an unintended pregnancy to term or had an abortion, and their partners. Participants were informed about the study at first consultation with their midwife, in the abortion clinic or via an online mailing directed at pregnant people. Because we did not want to interfere with the decision process, people could take part in the study after the abortion or when they had completed decision making and had decided to carry the pregnancy to term.

The RISE UP study (n = 262) included any person who was pregnancy and their partners, living in The Hague. Potential participants were informed about the study at their midwife’s practice, at Youth and Family Centres, via direct emails from an online pregnancy platform, and through social media advertisements. From April 2024 to October 2024, an additional test-retest sample of pregnant people in the RISE UP study (*n* = 110) was invited to complete the DA-LMUP again after 7 days.

#### 2.2.3 Procedure

For both studies, people could scan a QR-code that directed them to the information letter, video, and consent form. After receiving consent, the survey was sent. Upon completing the survey (about 20 minutes), they received a gift card worth €10. All survey items were obligatory and included answer options ‘I don’t know’ or ‘I’d rather not say’. For the test-retest sample, DA-LMUP items were not obligatory and did not include these two additional answer options.

#### 2.2.4 Measures

In addition to completing the DA-LMUP, participants reported on their age, and whether they currently had a romantic relationship. People who were (recently) pregnant were asked whether they were pregnant for the first time.

### 2.3 Analyses

Analyses were inspired by evaluations of the LMUP, hence both classical and modern test theory were applied (10, 12, 24, 25). As we developed two versions, for pregnant people and partners (Table 1), we performed analyses separately. All analyses were performed by MS & WB using R version 4.2.1. Answer options ‘I don’t know’ or ‘I’d rather not say’ were coded as missing. DA-LMUP data could be used if participants answered three or more questions. Mean imputation for remaining missing values was applied (26).

#### 2.3.1 Acceptability

To assess acceptability, we examined missing data rates with rates <5% indicating higher acceptability (27). Item discrimination was assessed by looking at item category endorsement values and checking if there were no item categories with endorsement (selection) >80%. We also looked at targeting by investigating if the distribution of our data covered the full range of the DA-LMUP, from 0 to 12. We investigated readability with the Flesch Reading Ease Score (FRES) and the Flesch-Kincaid Grade Level (FKGL) (28).

#### 2.3.2 Reliability

To assess reliability, we used the Cronbach’s alpha statistic with a score >0.7 indicating acceptable reliability (29). We also looked at corrected item-rest correlations (score <0.2 indicating limited contribution to homogeneity of the scale) and inter-item correlations (to check that all items had a positive direction). Additionally, we performed test-retest stability analyses using the weighted kappa and Pearson’s correlation – both acceptable if >0.60 (24, 30, 31). We also analysed the median and mean number of days between test and retest, and median and mean difference in the total DA-LMUP score.

#### 2.3.3 Construct validity

We assessed construct validity through a principal component analysis (PCA) to examine if all items were related to one construct, i.e., loading onto one component with an Eigenvalue of >1. Further, we performed a Confirmatory Factor Analysis (CFA) to investigate model fit, assessed by the Comparative Fit Index (CFI) (acceptable if > 0.95) and the Standardised Root Mean squared Residual (SRMR) (acceptable if <0.08) and Root Mean Squared Error Of Approximation (RMSEA) (acceptable if <0.06) (32).

Additionally, based on findings of previous studies, we tested the following hypotheses to examine whether the DA-LMUP behaved as theoretically expected: 1) people report a higher DA-LMUP score (i.e., more pregnancy intent) if they are continuing their pregnancy to term, compared to if people had an abortion (33, 34); 2) people who are in a relationship have higher DA-LMUP scores than people who are not in a relationship, with even higher scores for people who are cohabiting and/or married (12, 35-37); 3) people younger than 20 years or older than 39 years have lower DA-LMUP scores than those in between (34); 4) people who were pregnant for the first time have higher DA-LMUP scores than people who had been pregnant before (12, 35, 36, 38). Distribution of DA-LMUP scores and Shapiro-Wilk tests provided no evidence for normality (p<0.001 for both DA-LMUP versions), thus non-parametric Mann-Whitney U tests were used to test our hypotheses. For age, a Kruskal-Wallis test was performed, followed by a Dunn’s post-hoc test with a Bonferroni adjustment of the *p* values.

#### 2.3.4 Convergent validity

Data from couples were linked during recruitment, thus DA-LMUP scores could be compared through a paired samples t-test on couples’ difference scores. Further, the correlation between scores of partners was assessed. Although we expected a significant correlation between the scores, we did not expect perfect agreement (17). To test whether DA-LMUP scores differed between participants with and without a linked partner, a t-test was performed.

#### 2.3.5 Scaling

Mokken scale analysis tells us whether the separate items have different levels of difficulty. Participants should show as much agreement with items in line with the true extent of their pregnancy intention. The scalability of the Mokken scale was assessed by the Loevinger’s H coefficient, with only items of >0.3 eligible for scaling, while H<0.4 indicates a weak scale, 0.4<H<0.49 a medium scale, and ≥0.5 a strong scale (39).

## 3 Results

### 3.1 Sample

Of the 1201 participants, 1172 people (97.6%) answered 3 or more DA-LMUP items, and were therefore eligible to calculate a total DA-LMUP score. Of the total sample, 50.3% (n=590) were continuing their pregnancies, 20.6% (n=242) had recently had an abortion, 18.4% (n=216) were partners of those continuing a pregnancy, and 10.6% (n=124) were partners of those who had an abortion. Participants’ age ranged 16–55 years (people who were pregnant: median = 29, IQR = 25-33, range = 16-44 years; partners: median = 30, IQR = 25-33, range: 16-55 years). As shown in Table 2, most participants were born in the Netherlands, were not religious, and were cohabiting with a partner. Most pregnancies occurred within a relationship,53.2% of the pregnancies were first pregnancies (Table 2). Further, participants reported a broad range of pregnancy intent (Figure 1). Characteristics of the test-retest sample are reported in Supplementary Table 1.

**Table 2.** Socio-demographic characteristics of RISE UP and BluePrInt study participants (n = 1172).

| Characteristic | People who were pregnant |  |  | Partner |  |  |
| --- | --- | --- | --- | --- | --- | --- |
|  | Total<br>(n = 832)<br>n (%) | RISE UP<br>(n = 219)<br>n (%) | BluePrInt<br>(n = 613)<br>n (%) | Total<br>(n = 340)<br>n (%) | RISE UP<br>(n = 43)<br>n (%) | BluePrInt<br>(n = 297)<br>n (%) |
| Age groups |  |  |  |  |  |  |
| <20 | 37 (4.5) | 2 (0.9) | 35 (5.7) | 12 (3.5) | - | 12 (4.0) |
| 20-39 | 764 (91.8) | 209 (95.4) | 555 (90.5) | 308 (90.6) | 37 (86.0) | 271 (91.2) |
| >39 | 31 (3.7) | 8 (3.7) | 23 (3.8) | 20 (5.9) | 6 (14.0) | 14 (4.7) |
| Ethnicity |  |  |  |  |  |  |
| Born in the Netherlands,<br>both parents born in the<br>Netherlands | 657 (79.0) | 142 (64.8) | 515 (84.0) | 296 (87.1) | 28 (65.1) | 268 (90.2) |
| Born in the Netherlands,<br>one/both parents born<br>outside the Netherlands | 98 (11.8) | 41 (18.7) | 57 (9.3) | 29 (8.5) | 10 (23.3) | 19 (6.4) |
| Born outside the<br>Netherlands | 73 (8.8) | 36 (16.4) | 37 (6.0) | 15 (4.4) | 5 (11.6) | 10 (3.4) |
| Missing | 4 (0.5) | - | 4 (0.7) | - | - | - |
| Religion |  |  |  |  |  |  |
| No religion | 608 (73.1) | 144 (65.7) | 464 (75.7) | 282 (82.9) | 26 (60.5) | 256 (86.2) |
| Buddhism | 4 (0.5) | 2 (0.9) | 2 (0.3) | - | - | - |
| Christianity | 120 (14.4) | 31 (14.2) | 89 (14.5) | 31 (9.1) | 6 (14.0) | 25 (8.4) |
| Hinduism | 11 (1.3) | 7 (3.2) | 4 (0.7) | 1 (0.3) | 1 (2.3) | - |
| Islam | 47 (13.2) | 24 (11.0) | 23 (3.8) | 11 (3.2) | 5 (11.6) | 6 (2.0) |
| Jewish | - | - | - | 1 (0.3) | - | 1 (0.3) |
| Other | 18 (2.2) | 3 (1.4) | 15 (2.4) | 5 (1.5) | 2 (4.7) | 3 (1.0) |
| Missing | 24 (2.9) | 8 (3.6) | 16 (2.6) | 9 (2.7) | 3 (7.0) | 6 (2.0) |
| Partnership status |  |  |  |  |  |  |
| No relationship | 72 (8.7) | 9 (4.2) | 63 (10.3) | 17 (5.0) | - | 17 (5.7) |
| Not cohabiting with<br>partner | 139 (16.7) | 5 (2.3) | 134 (21.9) | 65 (19.1) | - | 65 (21.9) |
| Cohabiting with<br>partner | 611 (73.4) | 204 (93.2) | 407 (66.4) | 250 (73.5) | 43 (100.0) | 207 (69.7) |
| Missing | 10 (1.3) | 1 (0.5) | 9 (1.5) | 8 (2.2) | - | 8 (2.6) |
| Gestational age at survey |  |  |  |  |  |  |
| <4 weeks | 13 (1.6) | - | 13 (2.1) |  |  |  |
| 4-8 weeks | 185 (22.1) | 15 (6.8) | 170 (27.8) |  |  |  |
| 9-12 weeks | 190 (23.3) | 19 (8.7) | 171 (27.9) |  |  |  |
| 13-16 weeks | 242 (29.0) | 40 (18.3) | 202 (33.0) |  |  |  |
| 17-24 weeks | 102 (12.3) | 70 (32.0) | 32 (5.2) |  |  |  |
| >24 weeks | 92 (11.0) | 75 (34.2) | 17 (2.8) |  |  |  |
| Missing | 7 (0.8) | - | 7 (1.1) |  |  |  |
| Gravida |  |  |  |  |  |  |
| First pregnancy | 441 (53.1) | 93 (42.5) | 348 (56.9) |  |  |  |
| Second or subsequent<br>pregnancy | 380 (45.7) | 126 (57.5) | 254 (41.5) |  |  |  |
| Missing | 10 (1.2) | - | 10 (1.7) |  |  |  |

**Figure 1.**
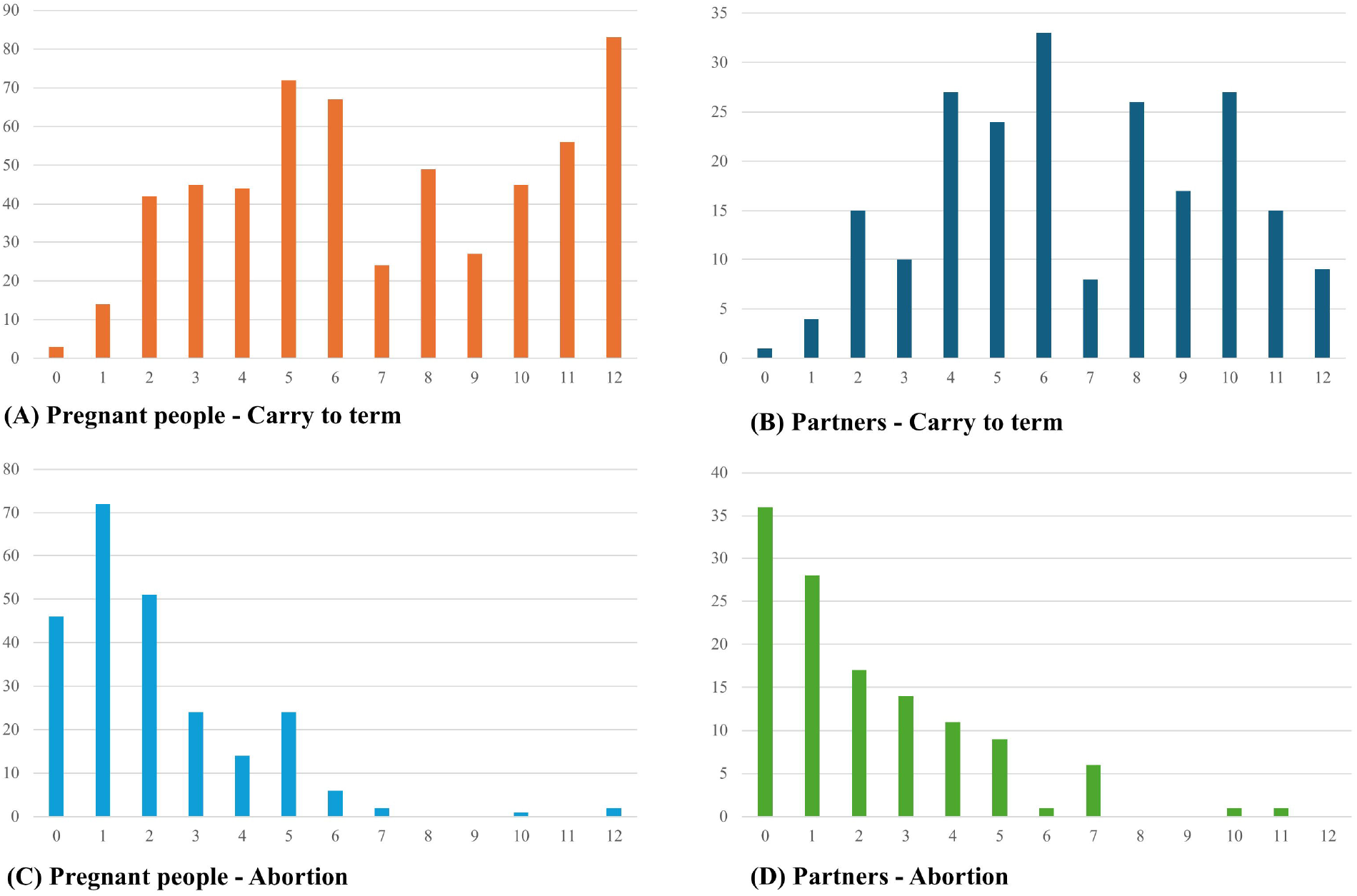
Distribution of the total DA-LMUP scores by DA-LMUP version and pregnancy decision.

### 3.2 Acceptability

Endorsement frequencies can be found in Supplementary Table 2. The number of missing responses per item ranged from 1.3% (item 1; contraception) to 16.7% (item 5; partner discussion) for people who were pregnant, and for partners from 1.2% (item 2; timing) to 23.5% (item 3; intention). Missing data rates of items 1, 2 and 6 were acceptable, while they were >5% for items 3, 4 and 5. Partners had more missing data than people who were pregnant for item 1 and 3. Participants did not have any missing data on item 6. Missing data was more common in the BluePrInt study compared to the RISE UP study. DA-LMUP scores ranged from 0 to 12 with a median of 5 and IQR from 2 to 8 (Figure 1), indicating a distribution of scores along the total scale. No question response had more than 80% endorsement on either version. For both versions, the FRES was 80 and the FKGL was 4.3.

### 3.3 Reliability

The Cronbach’s alpha indicated good internal consistency for people who were pregnant (0.88) and for partners (0.87). In the total sample, item-rest correlations were >0.2 for all items, and all inter-item correlations were positive (Table 3). As for the test-retest analyses (n = 101), mean time between test and retest was 9.3 (range 6-23 days) and the median difference was 0 (mean: 0.03). There were significant differences for ethnicity and gestational age between participants who did (n = 63) and who did not (n = 38) complete the retest. The weighted kappa and Pearson’s correlation values for individual items demonstrated good stability, with both scores ranging between 0.66 for item 5, and 0.91 for item 4. For the total score, weighted kappa was 0.52 and Pearson’s correlation coefficient was 0.97.

**Table 3.** Results of reliability, construct validity, and Mokken scale analyses of DA-LMUP for people who were pregnant and partners.

| Item | N | Item-rest correlation coefficient | PCA component loading | CFA factor loadings | Loevinger's H coefficient |
| --- | --- | --- | --- | --- | --- |
| People who were pregnant |  |  |  |  |  |
| Entire scale | 645 | Cronbach's alpha = 0.88 | Eigenvalue = 3.76 | CFI 0.930. SRMR 0.041.<br>RMSEA 0.160 (0.139-0.183) | 0.68 |
| 1 – Contraception | 821 | 0.57 | 0.28 | 0.45 | 0.46 |
| 2 – Timing | 817 | 0.78 | 0.44 | 0.82 | 0.69 |
| 3 – Intention | 778 | 0.86 | 0.45 | 0.84 | 0.78 |
| 4 – Desire | 721 | 0.85 | 0.45 | 0.86 | 0.72 |
| 5 – Partner discussion | 692 | 0.81 | 0.43 | 0.80 | 0.71 |
| 6 – Preparation | 832 | 0.67 | 0.38 | 0.65 | 0.68 |
| Partners |  |  |  |  |  |
| Entire scale | 226 | Cronbach's alpha = 0.87 | Eigenvalue = 3.76 | CFI 0.976. SRMR 0.035.<br>RMSEA 0.098 (0.059-0.139) | 0.68 |
| 1 – Contraception | 328 | 0.64 | 0.34 | 0.56 | 0.54 |
| 2 – Timing | 336 | 0.75 | 0.44 | 0.77 | 0.70 |
| 3 – Intention | 260 | 0.86 | 0.45 | 0.83 | 0.73 |
| 4 – Desire | 290 | 0.87 | 0.57 | 0.93 | 0.75 |
| 5 – Partner discussion | 248 | 0.87 | 0.46 | 0.90 | 0.75 |
| 6 – Preparation | 340 | 0.43 | 0.25 | 0.37 | 0.58 |

### 3.4 Construct validity

Table 3 presents results from PCA and CFA. PCA showed that all items loaded onto one dimension with an Eigenvalue of 3.76 both for people who were pregnant and partners. CFA moderately supported a single factor model for people who were pregnant with regards to CFI (0.930), SRMR (0.041), and RMSEA (0.160 (0.139-0.183)). A better fit was found for partners: CFI (0.976), SRMR (0.035), and RMSEA (0.098 (0.059-0.139)). All our hypothesis tests were confirmed (Table 4).

**Table 4.** Results from hypothesis testing. W values are presented for all tests except for age, there the values are a Kruskal-Wallis chi-squared statistic.

| **Hypothesis** | **People who were pregnant** |  | **Partners** |  |
| --- | --- | --- | --- | --- |
|  | **Test statistic** | **p-value** | **Test statistic** | **p-value** |
| Single vs Relationship | 17,953.0 | <0.001 | 1,670.0 | 0.009 |
| Non-cohabiting/single vs cohabiting | 32,328.0 | <0.001 | 4,302.5 | <0.001 |
| Age |  |  |  |  |
| 20 to 39 vs >39 | 2,540.0 | 0.021 | -2,320.0 | 0.030 |
| 20 to 39 vs <20 | 6,220.0 | <0.001 | 4,300.0 | <0.001 |
| >39 vs <20 | 2,450.0 | 0.021 | 4,930.0 | <0.001 |
| Abortion vs carry to term | 14,526.0 | <0.001 | 2,947.5 | <0.001 |
| Primigravida vs multigravida | 72,507.0 | <0.001 |  |  |

### 3.5 Convergent validity

Data were available for 257 couples. DA-LMUP scores of couples were highly correlated *(r* = 0.800). People who were pregnant reported lower pregnancy intentions compared to their partners (M_people who were pregnant_ = 4.48, M_partners_ = 4.94, *t*_*256*_ = -3.36, *p* <0.001).Additional results indicated that couple’s scores did not significantly differ in the abortion group (M_people who were pregnant, abortion_ = 1.70, M_partners, abortion_ = 1.94, *t*_*95*_ = -1.39, *p* = 0.167), though they did significantly differ in couples who continued the pregnancy to term (M_pregnant people, carry to term_ = 6.41, M_partners, carry to term_ = 6.73, *t*_*160*_ = -3.08, *p* = 0.002). Participants with a linked partner reported significantly lower pregnancy intentions than those without a linked partner (M_linked_ = 4.74, M_not linked_ = 6.10, *t*_*1169*_ = -6.38, *p* <0.001). Participants without a linked partner either did not have a partner or had a partner who did not take part.

### 3.6 Scaling

The Mokken scale analysis showed that items correspond to a basic Guttman structure with H>0.45 for all items for both versions (Table 3), meaning all items were eligible for scaling. For people who were pregnant, item 3 was easiest to endorse, followed by item 4, 2, 5, 6, and 1. For partners, item 4 was easiest to endorse, followed by 5, 3, 2, 6, and 1. Loevinger’s H was 0.68 for the entire scale for both versions, indicating a strong scale.

## 4 Discussion

In this study, we developed an acceptable and reliable Dutch Adaptation of the London Measure of Unplanned Pregnancy (DA-LMUP). The DA-LMUP was primarily based on the structure of the LMUP (10), with substantial linguistic and content-related modifications to enhance readability, acceptability, and cultural-linguistic appropriateness for the Dutch context. These included rewording items for clarity, adapting terminology, adding or modifying response options, and ensuring inclusivity by using gender-neutral or partner-specific language where relevant.

Overall, the DA-LMUP performed well according to various criteria for the performance of psychometric measures, both for people who were pregnant and their partners. The DA-LMUP appeared to be acceptable, readable and reliable. In construct validity analyses, all items loaded onto one dimension, moderately supporting a single factor model. Further, all hypotheses were confirmed. More specifically, this study shows that partners of people who have an abortion also report lower pregnancy intent. With regards to age, younger (<20 years) and older (>39 years) pregnant people reported lower pregnancy intent than those in between, aligning with a UK study (34), but contrasting with a Dutch study (38), which found higher pregnancy intentions in those over 39 years old. This inconsistency might be due to different study populations, as Maas et al. (38) focused on low-risk pregnancies continued to term. For partners, we found that a higher age was associated with higher pregnancy intent. Furthermore, results showed a high correlation of pregnancy intentions among couples, in line with previous studies.

Lastly, our Mokken Scale analyses indicated a strong scale. Notably, we found that item 1 (contraception) and item 6 (preparation behaviour) have the lowest contributions to the scale. This may reflect a broader disconnect between individuals’ behaviours and their underlying intentions: prior studies have identified discrepancies between reported pregnancy intentions and actual contraceptive practices (40, 41) and reported that some contraceptive behaviours, such as condom use or fertility-based methods, are not always perceived as ‘contraception’ (42). Similarly, although healthy preconception behaviours are widely promoted (43), even individuals who consciously plan their pregnancies often do not adhere to these recommendations (38, 44).

### 4.1 Strengths and limitations

This study has several strengths: we had a relatively large and diverse sample, containing data of both pregnant people and their partners, with a broad range of pregnancy intentions and different pregnancy outcomes (i.e., carry to term or abortion). Some limitations need to be acknowledged. First, the adjustments we made to the measure prevent direct comparison between the DA-LMUP and the LMUP. While these modifications were carefully considered and intended to better reflect the Dutch context, they inevitably altered the instrument. Second, people who were born outside the Netherlands, or with a parent born outside the Netherlands, and people with a religious background, were underrepresented in our sample (45, 46). Finally, some statistics (CFI for pregnant people and RMSEA for both) were (slightly) inadequate in the construct validity analyses.

### 4.2 Conclusion

The current study presents the DA-LMUP as a reliable instrument for assessing pregnancy intentions in research within the Dutch context. Future studies on pregnancy intention in the Netherlands are encouraged to apply the DA-LMUP to capture pregnancy intentions in a way that better reflects lived experiences compared to binary measures. This approach can provide policymakers with a better understanding of the diverse contexts and experiences surrounding pregnancy intentions in the Netherlands.

## Data Availability

All data produced in the present study are available upon reasonable request to the authors.

## Conflict of interest

The authors declare that the research was conducted in the absence of any commercial or financial relationships that could be construed as a potential conflict of interest.

## Author contributions

MS – Conceptualisation, Data curation, Formal analysis, Funding acquisition, Investigation, Methodology, Project administration, Writing – original draft

WB – Conceptualisation, Data curation, Formal analysis, Funding acquisition, Investigation, Methodology, Project administration, Writing – original draft

JvD – Conceptualisation, Funding acquisition, Methodology, Project administration, Supervision, Writing – review & editing

JKdJ – Conceptualisation, Funding acquisition, Methodology, Project administration, Supervision, Writing – review & editing

## Funding

Both studies were funded by ZonMw. The BluePrInt study under project number 554002012 and the RISE UP study in The Hague under project number 554002006.

## Acknowledgements

We would like to acknowledge all participants for responding to the survey and all people who have helped us to invite the population to participate in our studies. We want to thank the experienced experts and professional experts for their advice on the adaptations of the survey.

## Ethics statement

The requirement of ethical approval was waived by the Medical Research Ethics Committee of Leiden Den Haag Delft and the Medical Research Ethics Committee of the Amsterdam Medical Center for the studies involving humans because the Medical Research Involved Human subjects Act (WMO) did not apply. The studies were conducted in accordance with the local legislation and institutional requirements. The participants provided their written informed consent to participate in this study.

## Data availability statement

The datasets used and/or analysed during the current study are available from the corresponding author on reasonable request.

## Generative AI Statement

During the preparation of this work, the authors used ChatGPT in order to improve flow and conciseness of the content. After using this tool, the authors reviewed and edited the content as needed and take full responsibility for the content of the publication.

## Notes

### Competing Interest Statement

The authors have declared no competing interest.

### Funding Statement

This study was funded by ZonMw.

### Author Declarations

Ethics committees of Leiden Den Haag Delft and of the Amsterdam Medical Center waived ethical approval for this work.

### Summary of Updates

Following reviewer feedback, the instrument's name was changed to DA-LMUP and the manuscript's structure and conclusion were strengthened.

