## Supplementary material for "Pregnancy intentions in the Netherlands: An evaluation of a multidimensional and continuous construct": Surveys, Supplemental Table 1, Supplemental Table 2 (from another version of this preprint: version 3, 2025-08-05)

### 1 Supplementary Data

### 1.1 SPIN for pregnant people

*De volgende vragen gaan over de situatie en gevoelens in de periode rondom je huidige of meest recente zwangerschap.*

*Een voorbehoedsmiddel is iets wat je gebruikt om niet zwanger te worden. Bijvoorbeeld*

*de pil, een spiraal of condooms.*

1. **In de maand voordat ik zwanger werd gebruikte(n) ik/wij…**

*(geef aan welke stelling het best bij je past)*

- Geen voorbehoedsmiddelen
- Wel voorbehoedsmiddelen, maar niet altijd
- Altijd voorbehoedsmiddelen, maar weet nog dat het minstens één keer niet goed gewerkt heeft (bijv. condoom gescheurd, verschoven, pil uitgebraakt, etc.)
- Altijd voorbehoedsmiddelen en die hebben voor zover ik weet altijd goed gewerkt
- Weet ik niet
- Wens ik niet te beantwoorden

2. **Wat betreft ‘moeder/ouder worden’, heb ik het gevoel dat mijn zwangerschap plaatsvond op een…**

*(geef aan welke stelling het best bij je past)*

- Geschikt moment
- Oké moment, maar niet echt een geschikt moment
- Verkeerd moment
- Weet ik niet
- Wens ik niet te beantwoorden

3. **Net vóór ik zwanger werd…**

*(geef aan welke stelling het best bij je past)*

- Was ik van plan om zwanger te worden
- Veranderden mijn plannen om zwanger te worden steeds
- Was ik niet van plan om zwanger te worden
- Weet ik niet
- Wens ik niet te beantwoorden

4. **Net vóór ik zwanger werd…**

*(geef aan welke stelling het best bij je past)*

- Wilde ik een kind krijgen
- Twijfelde ik of ik een kind wilde krijgen
- Wilde ik geen kind krijgen
- Weet ik niet
- Wens ik niet te beantwoorden

*De volgende vraag gaat over jou en je partner/echtgenoot of degene van wie jij zwanger*

*bent.*

5. **Vóór ik zwanger werd…**

*(geef aan welke stelling het best bij je past)*

- Waren we het erover eens dat we (nog) een kind wilden krijgen
- Waren we het erover eens dat we geen kind (meer) wilden krijgen
- Hadden we het samen krijgen van kinderen besproken, maar waren we het niet eens
- Hadden we het samen krijgen van kinderen nooit besproken
- Heb ik ervoor gekozen om alleen zwanger te worden
- Weet ik niet
- Wens ik niet te beantwoorden

6. **Vóór je zwanger werd, heb je iets gedaan om je gezondheid te verbeteren als voorbereiding op de zwangerschap?**

*(je mag meerdere antwoorden aankruisen)*

- Ik nam foliumzuur
- Ik ben gestopt met of minder gaan roken
- Ik ben gestopt met of minder alcohol gaan drinken
- Ik ben gezonder gaan eten
- Ik ben meer gaan bewegen
- Ik zocht medisch of gezondheidsadvies, bijvoorbeeld bij de huisarts
- Ik heb iets anders gedaan, namelijk: ………………………………………………………………………………………
- Ik heb niets gedaan vóór mijn zwangerschap om mijn gezondheid te verbeteren
- Weet ik niet
- Wens ik niet te beantwoorden

### 1.2 SPIN for partners

*De volgende vragen gaan over de situatie en gevoelens in de periode rondom de huidige of meest recente zwangerschap.*

*Een voorbehoedsmiddel is iets wat je gebruikt om niet zwanger te worden. Bijvoorbeeld*

*de pil, een spiraal of condooms.*

1. **In de maand vóór de zwangerschap gebruikte(n) ik/wij…**

*(geef aan welke stelling het best bij je past)*

- Geen voorbehoedsmiddelen
- Wel voorbehoedsmiddelen, maar niet altijd
- Altijd voorbehoedsmiddelen, maar weet nog dat het minstens één keer niet goed gewerkt heeft (bijv. condoom gescheurd, verschoven, pil uitgebraakt, etc.)
- Altijd voorbehoedsmiddelen en die hebben voor zover ik weet altijd goed gewerkt
- Weet ik niet
- Wens ik niet te beantwoorden

2. **Wat betreft ‘vader/ouder worden’, heb ik het gevoel dat de zwangerschap plaatsvond op een…**

*(geef aan welke stelling het best bij je past)*

- Geschikt moment
- Oké moment, maar niet echt een geschikt moment
- Verkeerd moment
- Weet ik niet
- Wens ik niet te beantwoorden

3. **Net vóór de zwangerschap…**

*(geef aan welke stelling het best bij je past)*

- Was ik van plan om een kind te krijgen
- Veranderden mijn plannen om een kind te krijgen steeds
- Was ik niet van plan om een kind te krijgen
- Weet ik niet
- Wens ik niet te beantwoorden

4. **Net vóór de zwangerschap…**

*(geef aan welke stelling het best bij je past)*

- Wilde ik een kind krijgen
- Twijfelde ik of ik een kind wilde krijgen
- Wilde ik geen kind krijgen
- Weet ik niet
- Wens ik niet te beantwoorden

*De volgende vraag gaat over jou en je partner/echtgenoot of degene die van jou*

*zwanger is.*

5. **Vóór de zwangerschap…**

*(geef aan welke stelling het best bij je past)*

- Waren we het erover eens dat we (nog) een kind wilden krijgen
- Waren we het erover eens dat we geen kind (meer) wilden krijgen
- Hadden we het samen krijgen van kinderen besproken, maar waren we het niet eens
- Hadden we het samen krijgen van kinderen nooit besproken
- Weet ik niet
- Wens ik niet te beantwoorden

6. **Vóór de zwangerschap, heb je iets gedaan om je gezondheid te verbeteren als voorbereiding op de zwangerschap?**

*(je mag meerdere antwoorden aankruisen)*

- Ik ben gestopt met of minder gaan roken
- Ik ben gestopt met of minder alcohol gaan drinken
- Ik ben gezonder gaan eten
- Ik ben meer gaan bewegen
- Ik zocht medisch of gezondheidsadvies, bijvoorbeeld bij de huisarts
- Ik heb iets anders gedaan, namelijk: ………………………………………………………………………………………
- Ik heb niets gedaan vóór de zwangerschap om mijn gezondheid te verbeteren
- Weet ik niet
- Wens ik niet te beantwoorden

### 2 Supplementary Tables

### Supplementary Table 1. Socio-demographic characteristics of the test-retest sample.

| **Characteristic** | **Survey (n = 101)**  **n (%)** | **Retest (n = 63)**  **n (%)** | **Non-retest(n = 38)**  **n (%)** | **Retest**  **versus non-retest** |
| --- | --- | --- | --- | --- |
| Age groups |  |  |  |  |
| <20 | 0 (0.0) | 0 (0.0) | 0 (0.0) | p = 0.630 |
| 20-39 | 97 (96) | 61 (96.8) | 36 (94.7) |  |
| >39 | 4 (4) | 2 (3.2) | 2 (5.3) |  |
| Ethnicity |  |  |  |  |
| Born in the Netherlands | 58 (57.4) | 44 (69.8) | 14 (36.8) | p = 0.004 |
| One or both parents born outside the Netherlands | 28 (27.7) | 13 (20.6) | 15 (39.5) |  |
| Born outside the Netherlands | 15 (14.9) | 6 (9.5) | 9 (23.7) |  |
| Missing | 0 (0.0) | 0 (0.0) | 0 (0.0) |  |
| Religion |  |  |  |  |
| No religion | 56 (55.4) | 38 (60.3) | 18 (47.4) | p = 0.051 |
| Buddhism | 0 (0.0) | 0 (0.0) | 0 (0.0) |  |
| Christianity | 15 (14.9) | 11 (17.5) | 4 (10.5) |  |
| Hinduism | 4 (4) | 0 (0.0) | 4 (10.5) |  |
| Islam | 18 (17.8) | 8 (12.7) | 10 (26.3) |  |
| Jewish | 0 (0.0) | 0 (0.0) | 0 (0.0) |  |
| Other | 3 (3) | 2 (3.2) | 1 (2.6) |  |
| Missing | 5 (5) | 4 (6.4) | 1 (2.6) |  |
| Partnership status |  |  |  |  |
| No relationship | 6 (5.9) | 2 (3.2) | 4 (10.5) | p = 0.196 |
| Not cohabiting with partner | 7 (6.9) | 3 (4.8) | 4 (10.5) |  |
| Cohabiting with partner | 87 (86.1) | 57 (90.5) | 30 (78.9) |  |
| Missing | 1 (1) | 1 (1.6) | 0 (0.0) |  |
| Gestational age at survey |  |  |  |  |
| <4 weeks | 0 (0.0) | 0 (0.0) | 0 (0.0) | p = 0.020 |
| 4-8 weeks | 0 (0.0) | 0 (0.0) | 0 (0.0) |  |
| 9-12 weeks | 11 (11.6) | 6 (9.8) | 5 (14.7) |  |
| 13-16 weeks | 9 (9.5) | 8 (13.1) | 1 (2.9) |  |
| 17-24 weeks | 33 (34.7) | 26 (42.6) | 7 (20.6) |  |
| >24 weeks | 42 (44.2) | 21 (34.4) | 21 (61.8) |  |
| Missing | 0 (0.0) | 0 (0.0) | 0 (0.0) |  |
| Gravida |  |  |  |  |
| First pregnancy | 36 (35.6) | 22 (34.9) | 14 (36.8) | p = 1.00 |
| Second or subsequent pregnancy | 65 (64.4) | 41 (65.1) | 24 (63.2) |  |
| Missing | 0 (0.0) | 0 (0.0) | 0 (0.0) |  |

NB: Characteristics are presented of participants who completed all LMUP items (n = 101).

### Supplementary Table 2. Endorsement frequencies of LMUP items and response options (n = 1172).

|  | **People who were pregnant** | | | **Partners** | | |
| --- | --- | --- | --- | --- | --- | --- |
|  | **All**  **(n = 832)** | **Decision:**  **Carry to term**  **(n = 590)** | **Decision:**  **Abortion**  **(n = 242)** | **All**  **(n = 340)** | **Decision:**  **Carry to term**  **(n = 216)** | **Decision: Abortion**  **(n = 124)** |
| **Item** | **N** **(%)** | **N** **(%)** | **N** **(%)** | **N** **(%)** | **N** **(%)** | **N** **(%)** |
| 1 – Contraception |  |  |  |  |  |  |
| Always using contraception | 161 (19.4) | 77 (13.1) | 84 (34.7) | 90 (26.5) | 31 (14.4) | 59 (47.6) |
| Using sometimes or failed at least once | 281 (33.8) | 184 (31.2) | 97 (40.1) | 124 (36.5) | 77 (35.7) | 47 (37.9) |
| Not using contraception | 379 (45.6) | 318 (53.9) | 61 (25.2) | 114 (33.5) | 96 (44.4) | 18 (14.5) |
| *Don't know* | 4 (0.5) | 4 (0.7) | 0 (0.0) | 0 (0.0) | 12 (5.6) | 0 (0.0) |
| *Rather not say* | 7 (0.8) | 7 (1.2) | 0 (0.0) | 12 (3.6) | 0 (0.0) | 0 (0.0) |
| *Missing* |  |  |  |  |  |  |
| 2 – Timing |  |  |  |  |  |  |
| Wrong time | 257 (30.9) | 68 (11.5) | 189 (78.1) | 97 (28.5) | 15 (6.9) | 82 (66.1) |
| OK time | 314 (37.7) | 274 (46.4) | 40 (16.5) | 142 (41.8) | 109 (50.5) | 33 (26.6) |
| Right time | 246 (29.6) | 241 (40.8) | 5 (2.1) | 97 (28.5) | 91 (42.1) | 6 (4.8) |
| *Don't know* | 11 (1.3) | 5 (0.9) | 6 (2.5) | 4 (1.2) | 1 (0.5) | 3 (2.4) |
| *Rather not say* | 4 (0.5) | 2 (0.3) | 2 (0.8) | 0 (0.0) | 0 (0.0) | 0 (0.0) |
| *Missing* |  |  |  |  |  |  |
| 3 – Intention |  |  |  |  |  |  |
| Did not intend pregnancy | 481 (57.8) | 261 (44.2) | 220 (90.9) | 143 (42.1) | 45 (20.8) | 98 (79.0) |
| Intentions kept changing | 98 (11.8) | 84 (14.2) | 14 (5.8) | 46 (13.5) | 30 (13.9) | 16 (12.9) |
| Intended pregnancy | 199 (23.9) | 196 (33.2) | 3 (1.2) | 71 (20.9) | 68 (31.5) | 3 (2.4) |
| *Don't know* | 42 (5) | 38 (6.4) | 4 (1.7) | 64 (18.8) | 57 (26.4) | 7 (5.6) |
| *Rather not say* | 12 (1.4) | 11 (1.9) | 1 (0.4) | 16 (4.7) | 16 (7.4) | 0 (0.0) |
| *Missing* |  |  |  |  |  |  |
| 4 – Desire |  |  |  |  |  |  |
| Did not want a baby | 242 (29.1) | 62 (10.5) | 180 (74.4) | 99 (29.1) | 14 (6.5) | 85 (68.6) |
| Mixed feelings about having a baby | 168 (20.2) | 131 (22.2) | 37 (15.3) | 94 (27.7) | 66 (30.6) | 28 (22.6) |
| Wanted baby | 199 (23.9) | 291 (49.3) | 20 (8.3) | 97 (28.5) | 89 (41.2) | 8 (6.4) |
| *Don't know* | 89 (10.7) | 84 (14.3) | 5 (2.1) | 40 (11.8) | 37 (17.1) | 3 (2.4) |
| *Rather not say* | 22 (2.6) | 22 (3.7) | 0 (0.0) | 10 (2.9) | 10 (4.6) | 0 (0.0) |
| *Missing* |  |  |  |  |  |  |
| 5 – Partner discussion |  |  |  |  |  |  |
| Never discussed getting pregnant with partner, or agreed to not get pregnant | 242 (29.1) | 80 (13.6) | 162 (66.9) | 109 (32.1) | 25 (11.6) | 84 (67.7) |
| Discussed, but did not agree about getting pregnant | 78 (9.4) | 47 (8.0) | 31 (12.8) | 36 (10.6) | 15 (6.9) | 21 (16.9) |
| Agreed to get pregnant | 372 (44.8) | 342 (58.1) | 30 (12.4) | 103 (30.3) | 90 (41.7) | 13 (10.5) |
| *Don't know* | 105 (12.6) | 89 (15.1) | 16 (6.6) | 65 (19.1) | 59 (27.3) | 6 (4.8) |
| *Rather not say* | 34 (4.1) | 31 (5.3) | 3 (1.2) | 27 (7.9) | 27 (12.5) | 0 (0.0) |
| *Missing* |  |  |  |  |  |  |
| 6 – Preparation |  |  |  |  |  |  |
| No preparatory lifestyle changes | 532 (63.9) | 314 (53.2) | 218 (90.1) | 266 (78.2) | 150 (69.4) | 116 (93.6) |
| Did 1 preparatory lifestyle change | 144 (17.3) | 129 (21.9) | 15 (6.2) | 43 (12.7) | 38 (17.6) | 5 (4.0) |
| Did 2 or more preparatory lifestyle changes | 156 (18.8) | 147 (24.9) | 9 (3.7) | 31 (9.1) | 28 (13.0) | 3 (2.4) |
| *Don't know* | 0 (0.0) | 0 (0.0) | 0 (0.0) | 0 (0.0) | 0 (0.0) | 0 (0.0) |
| *Rather not say* | 0 (0.0) | 0 (0.0) | 0 (0.0) | 0 (0.0) | 0 (0.0) | 0 (0.0) |
| *Missing* |  |  |  |  |  |  |
